# SARS-CoV-2-reactive interferon-γ-producing CD8^+^ T cells in patients hospitalized with Coronavirus viral disease-2019

**DOI:** 10.1101/2020.05.18.20106245

**Authors:** Estela Giménez, Eliseo Albert, Ignacio Torres, María José Remigia, María Jesús Alcaraz, María José Galindo, María Luisa Blasco, Carlos Solano, María José Forner, Josep Redón, Jaime Signes-Costa, David Navarro

**Affiliations:** Microbiology Service, Clinic University Hospital, INCLIVA Health Research Institute, Valencia, Spain.; Hematology Service, Clinic University Hospital, INCLIVA Health Research Institute, Valencia, Spain.; Internal Medicine Department, Clinic University Hospital, INCLIVA Health Research Institute, Valencia, Spain.; Medical Intensive Care Unit, Clinic University Hospital, INCLIVA Health Research Institute, Valencia, Spain.; Department of Medicine, School of Medicine, University of Valencia, Valencia, Spain.; Pneumology Service, Clinic University Hospital, INCLIVA Health Research Institute, Valencia, Spain; Department of Microbiology, School of Medicine, University of Valencia, Valencia, Spain.

**Keywords:** SARS-CoV-2, Covid-19, CD8^+^ T cells, T-cell immunity

## Abstract

There is limited information on SARS-CoV-2 T-cell immune responses in patients with Covid-19. Both CD4^+^ and CD8^+^ T cells may be instrumental in the resolution of and protection from SARS-CoV-2 infection. Here, we tested 25 hospitalized patients with either microbiologically documented Covid-19 (n=19) or highly suspected of having the disease (n=6) for the presence of SARS-CoV-2-reactive-CD69^+^-expressing interferon--producing-(IFN-γ) CD8^+^ T cells by a flow-cytometry for intracelular cytokine staining assay. Two sets of overlapping peptides encompassing the SARS-CoV-2 Spike glycoprotein N-terminal 1-643 amino acid sequence and the entire sequence of SARS-CoV-2 M protein were used simultaneously as antigenic stimulus. Ten patients (40%) had detectable responses, displaying frequencies ranging from 0.15 to 2.7% (median of 0.57 cells/μL; range, 0.43-9.98 cells/μL). The detection rate of SARS-CoV-2-reactive IFN-γ CD8^+^ T cells in patients admitted to intensive care was comparable (*P*=0.28) to that in patients hospitalized in other medical wards. No correlation was found between SARS-CoV-2-reactive IFN-γ CD8^+^ T-cell counts and SARS-CoV-2 S-specific antibody levels. Likewise, no correlation was observed between either SARS-CoV-2-reactive IFN-γ CD8^+^ T cells or S-specific IgG-antibody titers and blood cell count or levels of inflammatory biomarkers. In summary, in this descriptive, preliminary study we showed that SARS-CoV-2-reactive IFN-γ CD8^+^ T cells can be detected in a non-negligible percentage of patients with moderate to severe forms of Covid-19. Further studies are warranted to determine whether quantitation of these T-cell subsets may provide prognostic information on the clinical course of Covid-19.

## INTRODUCTION

On March 11, 2020 the World Health Organization (WHO) declared Covid-19 a pandemic^1^. As of May 12^th^ more than 4,200,000 cases of Covid-19 have been reported worldwide, causing over 290,000 deaths.^2^ Covid-19 commonly results in pneumonia, which could evolve to acute respiratory distress syndrome, leading to respiratory or multiorgan failure^3,4^. Elucidation of immune responses conferring protection against SARS-CoV-2 is crucial to develop an effective vaccine prototype, which is urgently needed to blunt pandemic progression. A number of studies have focused on the characterization of SARS-CoV-2-specific antibody kinetics profiles^5-10^. Nevertheless, there is scant information on T-cell responses against SARS-CoV-2 in Covid-19 patients. CD4^+^ and CD8^+^ T cells targeting structural viral proteins proteins appear to confer broad and long-lasting protection against SARS-CoV^11^. Several clusters of cytotoxic T lymphocyte (CTL) epitopes restricted by HLA-class I specificities commonly found in caucasians (i.e. HLA-A02) have been mapped within the spike (S) and the membrane (M) SARS-CoV proteins.^11^ Since SARS-CoV shares high sequence identity with SARS-CoV-2,^12^ it is reasonable to expect an analogous scenario in the SARS-CoV-2 infection. Braun and colleagues demonstrated the presence of SARS-CoV-2-S-reactive, activated (expressing CD38^+^ and HLA-DR) CD4^+^ T cells in 83% of Covid-19 patients.^13^ No data on the potential functionality of these cells was provided.^13^ Ni et al. detected T-cell responses against several SARS-CoV-2 structural proteins, as measured by interferón-y ELISPOT, in up to 50% of Covid-19 convalescent patients.^14^ Here, we developed a flow-cytometry for intracelular cytokine staining (ICS) assay to enumerate peripheral blood SARS-CoV-2-reactive-IFN-γ-producing CD8^+^ T cells, which was used to assess the presence of virus-elicited T-cell immunity in patients with moderate to severe Covid-19.

## PATIENTS AND METHODS

### Patients

Twenty-five non-consecutive patients (14 males and 11 females; median age, 69 years; range, 62 to 87 years) admitted to our center from March 17^th^ to April 24^th^ with clinically suspected Covid-19 were included in the current study. The only inclusion criterion was the availability of blood specimens for T-cell immunity analyses. Medical history and laboratory data were retrospectively reviewed. The current study was approved by the Ethics Comittee of Hospital Clínico Universitario INCLIVA.

### Detection of SARS-CoV-2 by RT-PCR

Nasopharyngeal or oropharygeal specimens were obtained with flocked swabs in universal transport medium (Beckton Dickinson, Sparks, MD, USA, or Copan Diagnostics, Murrieta, CA, USA) and conserved at 4 °C until processed (within 6 hours). Tracheal aspirates from mechanically ventilated patients were also processed, when available. These latter specimens were collected undiluted. Nucleic acid extraction was performed using the Qiagen EZ-1 Viral extraction kit or the DSP virus Pathogen Minikit on the EZ1 or QiaSymphony Robot instruments (Qiagen, Valencia, CA, USA), respectively. Commercially-available RT-PCR assays used for SARS-CoV-2 testing were one or more of the following: E-gene/LightMix® Modular SARS-CoV-2 (COVID-19) RdRP gene from TIB MOLBIOL GmHD, distributed by Roche Diagnostics (Pleasenton, CA, USA) on the Light Cycler 2.0 instrument, the SARS-COV-2 REALTIME PCR KIT from Vircell Diagnostics (Granada, Spain), the REALQUALITY RQ-2019-nCoV from AB ANALITICA (Padua, Italy), both on the Applied Biosystems 7500 instrument, the SARS-CoV-2 (S gene) – BD MAX™ System (VIASURE Real Time PCR Detection Kits; CerTest, Zaragoza, Spain) and the Abbott RealTime SARS-CoV-2 Assay (Abbott Molecular Diagnostics, Chicago, USA). A number of these specimens were screened for the presence of other respiratory pathogens by the the NxTAG® Respiratory Pathogen Panel (Luminex Corp, Austin, Tx, USA).

### Antibody detection methods

Initial screening for SARS-CoV-2-specific antibodies was carried out by using the 2019-nCoV IgG/IgM Rapid Test (Hangzhou AllTest Biotech Co., Ltd. China), a rapid lateral flow chromatographic immunoassay (LFIC) designed for the qualitative detection of IgG and IgM antibodies in human whole blood or serum. Sera obtained at the time of blood collection for SARS-CoV-2 CD8^+^ T cell immunity analyses were analyzed by the LIAISON® SARS-CoV-2 IgG (DiaSorin, Saluggia, Italy), a fully automated quantitative chemiluminscent assay (CLIA) detecting IgG antibodies against SARS-CoV-2 S protein (S1 and S2 subunits). Immunoassays were performed and interpreted according the instructions of the respective manufacturer.

### SARS-CoV-2-reactive IFN-γ CD8^+^ T cells

Enumeration of SARS-CoV-2-reactive CD69^+^-expressing-IFNγ-producing-CD8^+^ T cells was carried out by flow cytometry for ICS (BD Fastimmune, BD-Beckton Dickinson and Company-Biosciences, San Jose, CA, USA), following an adapted protocol developed by our group for quantitation of Cytomegalovirus-specific T cells.^15,16^ Briefly, heparinized whole blood (0.5 ml) was simultaneously stimulated for 6 h with two sets of 15-mer overlapping peptides (11 mer overlap) encompassing the SARS-CoV-2 Spike glycoprotein N-terminal 1-643 amino acid sequence (158 peptided) and the entire sequence of SARS-CoV-2 M protein (53 peptides), at a concentration of 1 μg/ml per peptide, in the presence of 1 μg/ml of costimulatory mAbs to CD28 and CD49d. Peptide mixes were obtained from JPT peptide Technologies GmbH (Berlin, Germany). Samples mock stimulated with PBS/DMSO and costimulatory antibodies were run in parallel. Brefeldin A (10 μg/ml) was added for the last 4 h of incubation. Blood was then lysed (BD FACS lysing solution) and frozen at −80 °C until tested. On the day of testing, stimulated blood was thawed at 37 J°C, washed, permeabilized (BD permeabilizing solution), and stained with a combination of labeled moAbs (anti-IFNγ-FITC, anti-CD69-PE, anti-CD8-PerCP-Cy5.5, and anti-CD3-APC) for 1 h at room temperature. Appropriate positive (phytohemagglutinin) and isotype controls were used. Cells were then washed, resuspended in 200 μl of 1% paraformaldehyde in PBS, and analyzed within 2 h on an FACScanto flow cytometer using DIVA v8 software (BD Biosciences Immunocytometry Systems, San Jose, CA, USA). CD3/CD8^+^ events were gated and then analyzed for the CD69^+^ activation marker and IFNγ production (Figure 1A). The total number of SARS-CoV-2-reactive CD8^+^ T cells was calculated by multiplying the percentages of CD8^+^ T cells producing IFNγ on stimulation (after background subtraction) by the absolute CD8^+^ T cell count. Responses ≥0.1% were considered specific.

**Figure 1.**
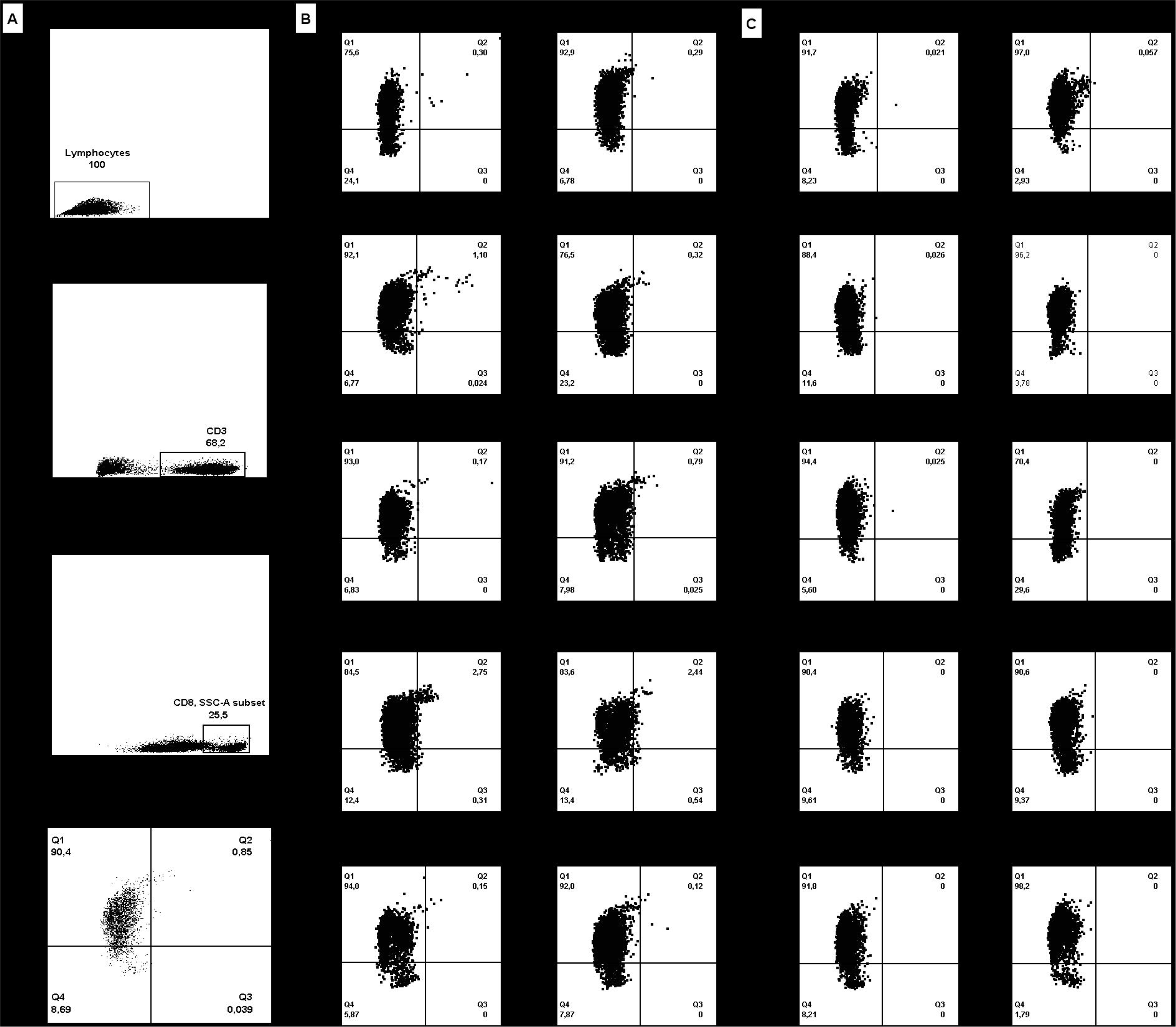
Enumeration of SARS-CoV-2-S1/M-reactive CD69^+^-expressing IFN-γ-producing CD8^+^ T cells by flow cytometry for intracellular staining in Covid-19 patients. Panel A depicts the gating strategy. Panel B includes plots from patients with detectable responses, while in panel C plots from 10 patients testing negative are shown. Dot-plot figures were built with FlowIo software (BD Biosciences).

### Laboratory measurements

Clinical laboratory investigation included a complete blood count and serum levels of ferritin, Dimer-D, and C reactive protein (CRP). Data on serum IL-6 levels were not available.

### Statistical methods

Frequency comparisons for categorical variables were carried out using the Fisher exact test. Differences between medians were compared using the Mann–Whitney U-test. The Spearman’s rank test was used for analysis of correlation between continuous variables.

Two-sided exact *P*-values were reported. A *P*-value <0.05 was considered statistically significant. The analyses were performed using SPSS version 20.0 (SPSS, Chicago, IL, USA).

## RESULTS

### Patient clinical features

Patients in this cohort were admitted to our center at a median of 7 days (range, 0-28 days) since the onset of symptoms. Twenty-two patients presented with pneumonia and imaging findings on chest-x ray or CT-scans compatible with Covid-19. The remaining three patients, clinically suspected of Covid-19 with no evidence of pneumonia, were admitted either due to aggravation of baseline chronic conditions (n=2) or venous thrombosis. Median hospitalization time of patients was 18 days (range, 4-52 days). Seven patients needed intensive care (ICU), of whom 2 died.

As shown in Table 1, diagnosis of SARS-CoV-2 was achieved in 17 patients by RT-PCR in upper or lower respiratory tract specimens, either at initial screening (n=10) or after repeat testing (n=7). Of these, 14 also had serological evidence of SARS-CoV-2 infection. Two patients tested repeatedly negative by RT-PCR, but exhibited IgG seroconversion. Finally, microbiological evidence of SARS-CoV-2 infection could no be obtained in 6 patients. These latter patients tested negative by a multiplexed RT-PCR assay targeting prevalent respiratory viruses and bacteria in upper or lower respiratory tract specimens.

**Table 1.**
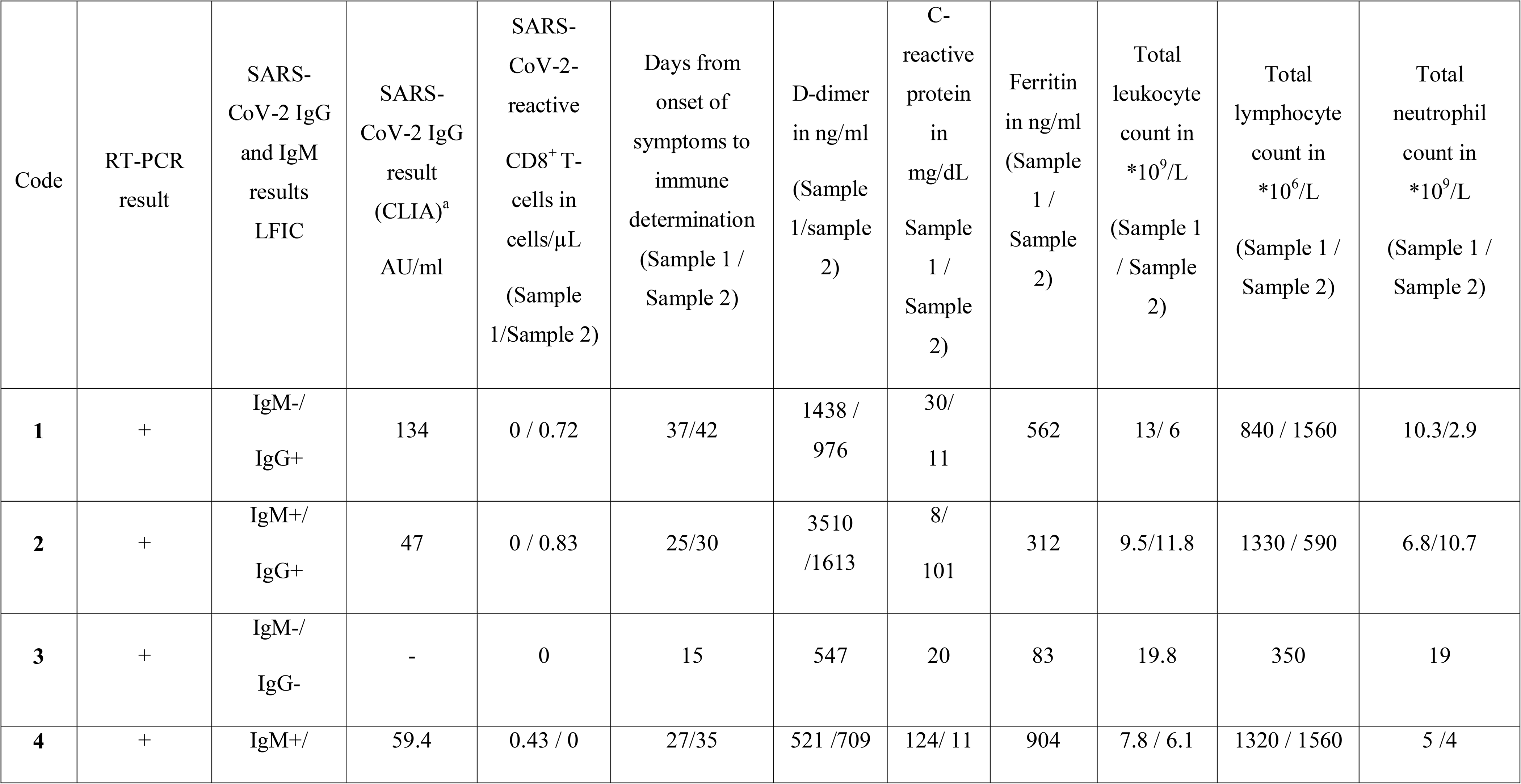
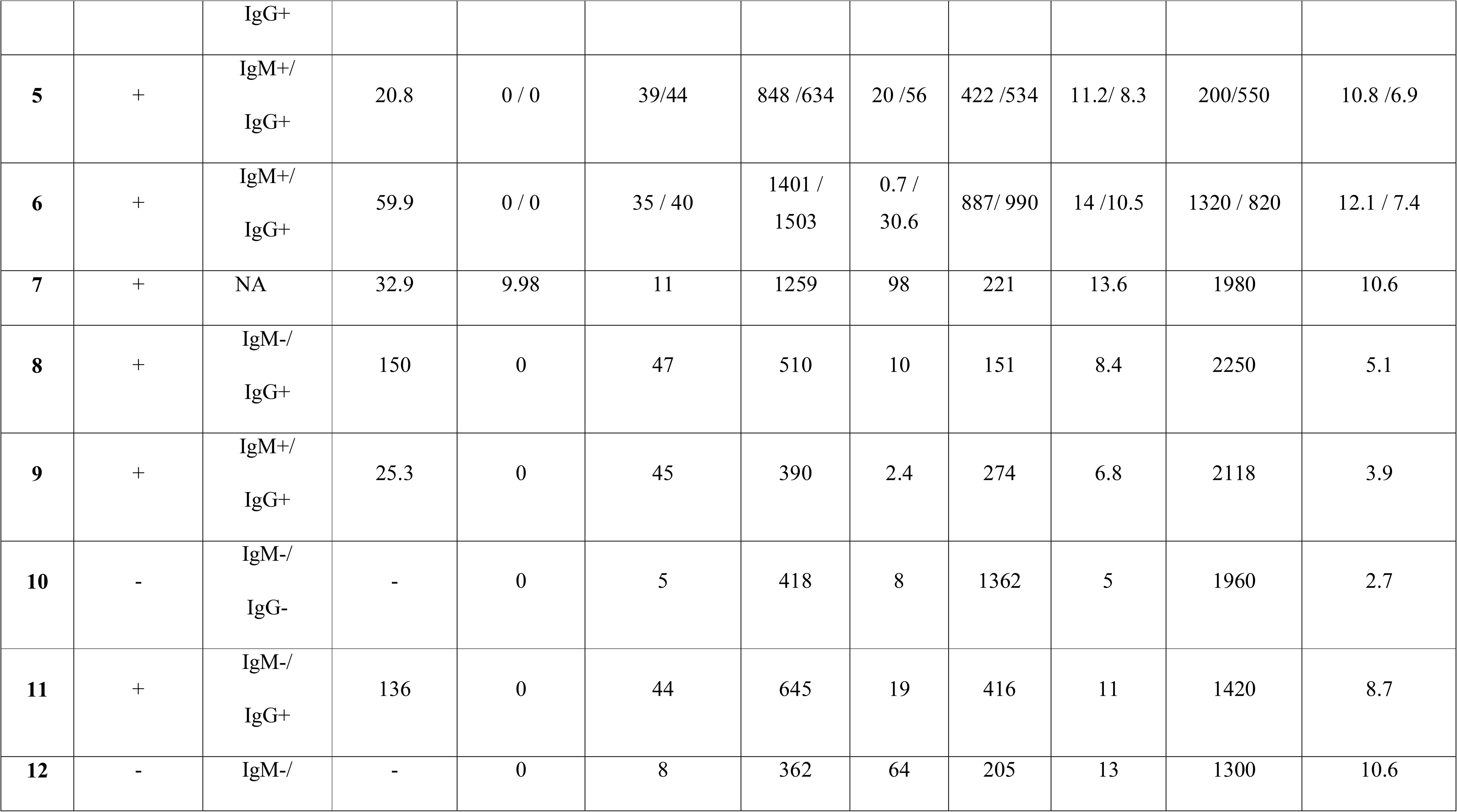
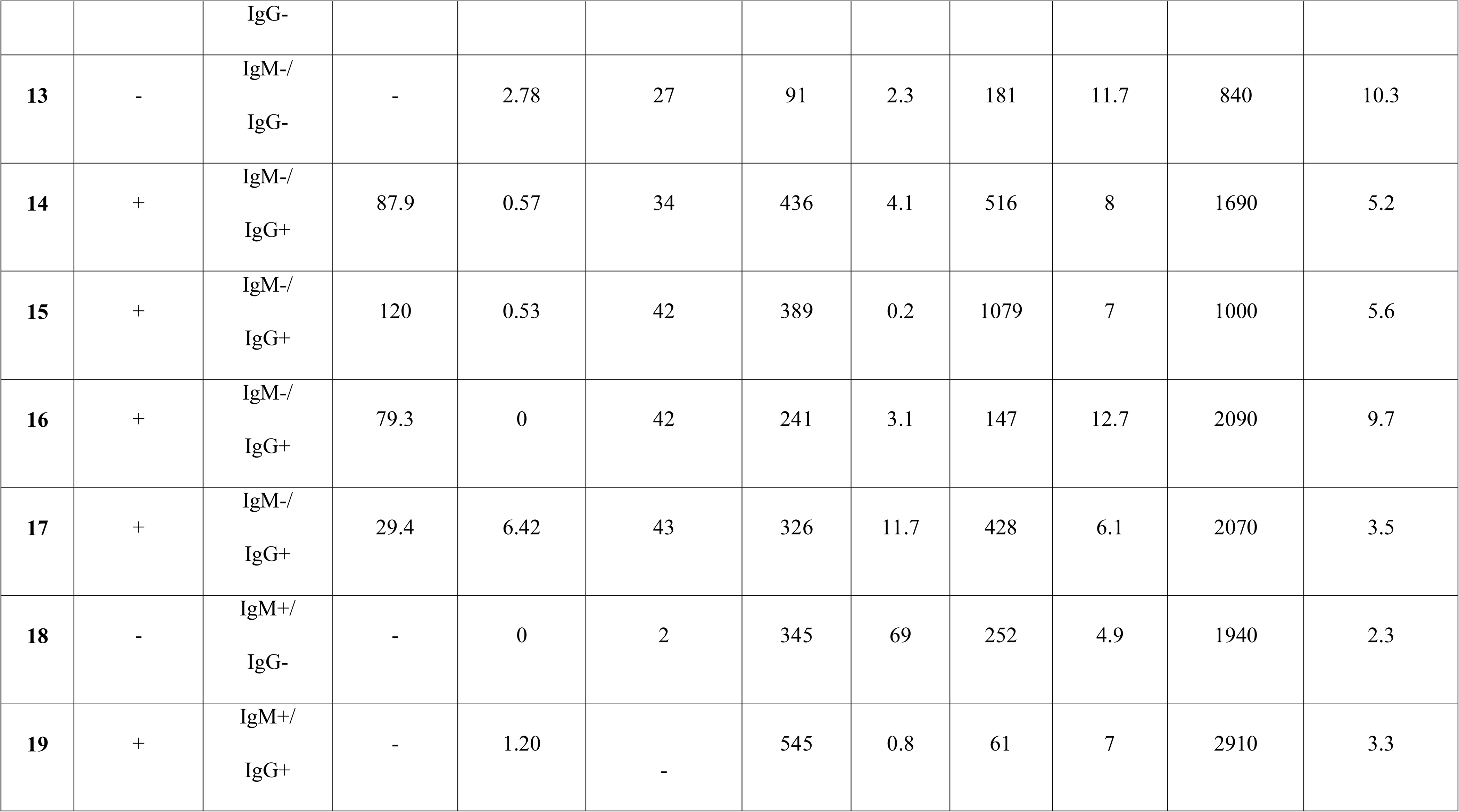
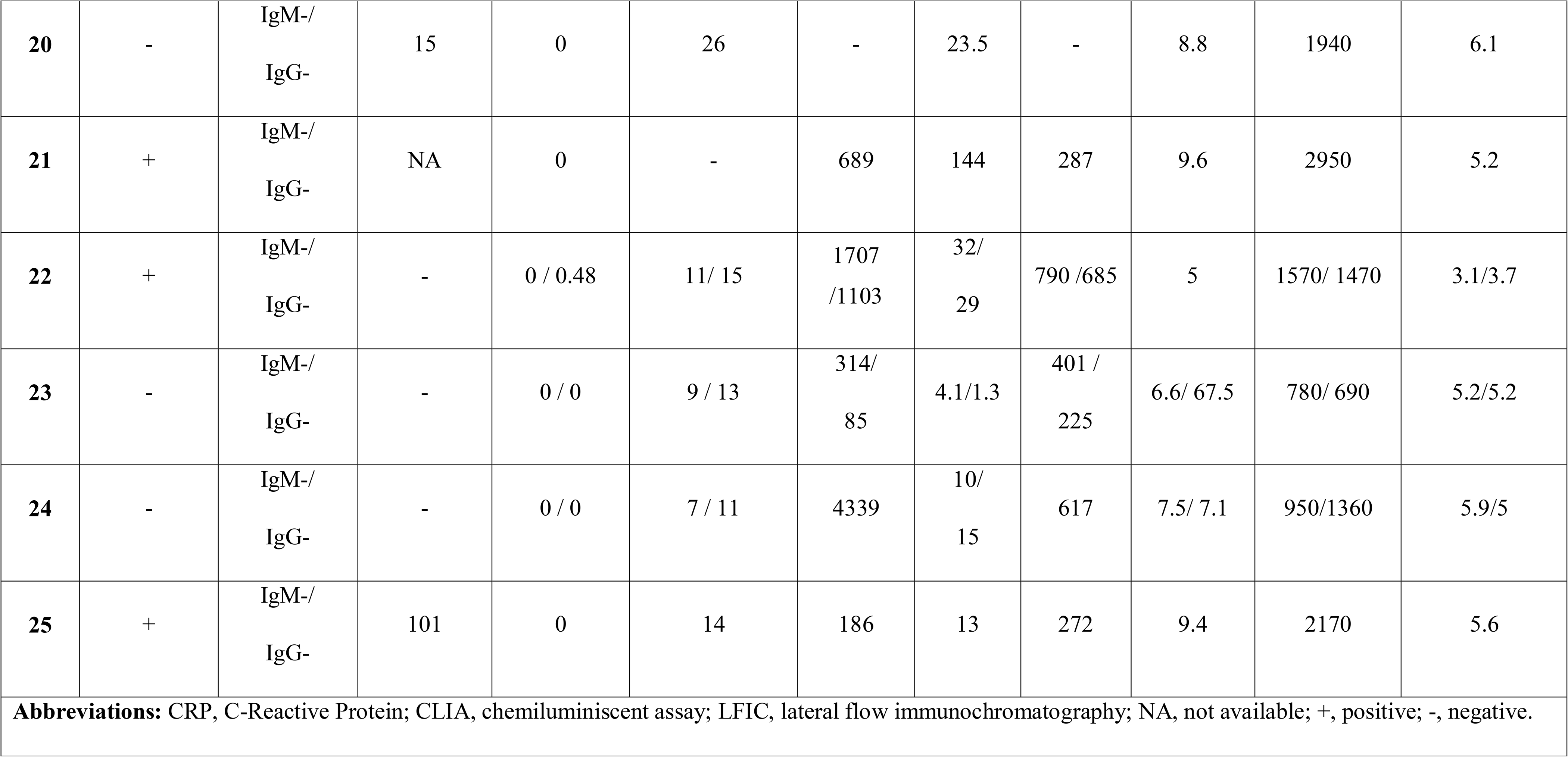
Microbiological and laboratory data on patients with Covid-19 included in the study.

### Detection of SARS-CoV-2-reactive IFN-γ CD8^+^ T cells in Covid-19 patients

Enumeration of peripheral blood SARS-CoV-2-S1/M-reactive CD69^+^-IFN-γ CD8^+^ T cells was carried out at a median of 27 days from the onset of symptoms (range, 2-47 days). Eight patients were re-screened within the next 5 days. Ten patients (40%) had detectable responses (Table 1 and Figure 1B), displaying frequencies ranging from 0.15 to 2.7% (median of 0.57 cells/μL; range, 0.43-9.98 cells/μL). Nine of these patients had previously tested positive by RT-PCR, and 8 had specific antibodies detected by LFIC, CLIA, or both. Among the latter patients, 3 had IgM antibodies. A single patient (number 13 in Table 1) displayed detectable SARS-CoV-2-reactive IFN-γ CD8^+^ T cells despite a lack of microbiological confirmation of Covid-19 by either RT-PCR or serological methods. Fifteen pateints had no detectable SARS-CoV-2 IFNγ CD8^+^ T-cell responses (representative examples shown in Figure 1C).

The detection rate of SARS-CoV-2-reactive IFN-γ CD8^+^T cells in patients admitted to ICU (57%) was not significantly different *P*=0.28) from that in patients hospitalized in other medical wards (33%).

No correlation was found between levels of SARS-CoV-2-reactive IFN-γ CD8^+^ T cells and those of SARS-CoV-2 S-specific antibodies quantitated by CLIA (Figure 2) in paired whole blood and serum specimens.

**Figure 2.**
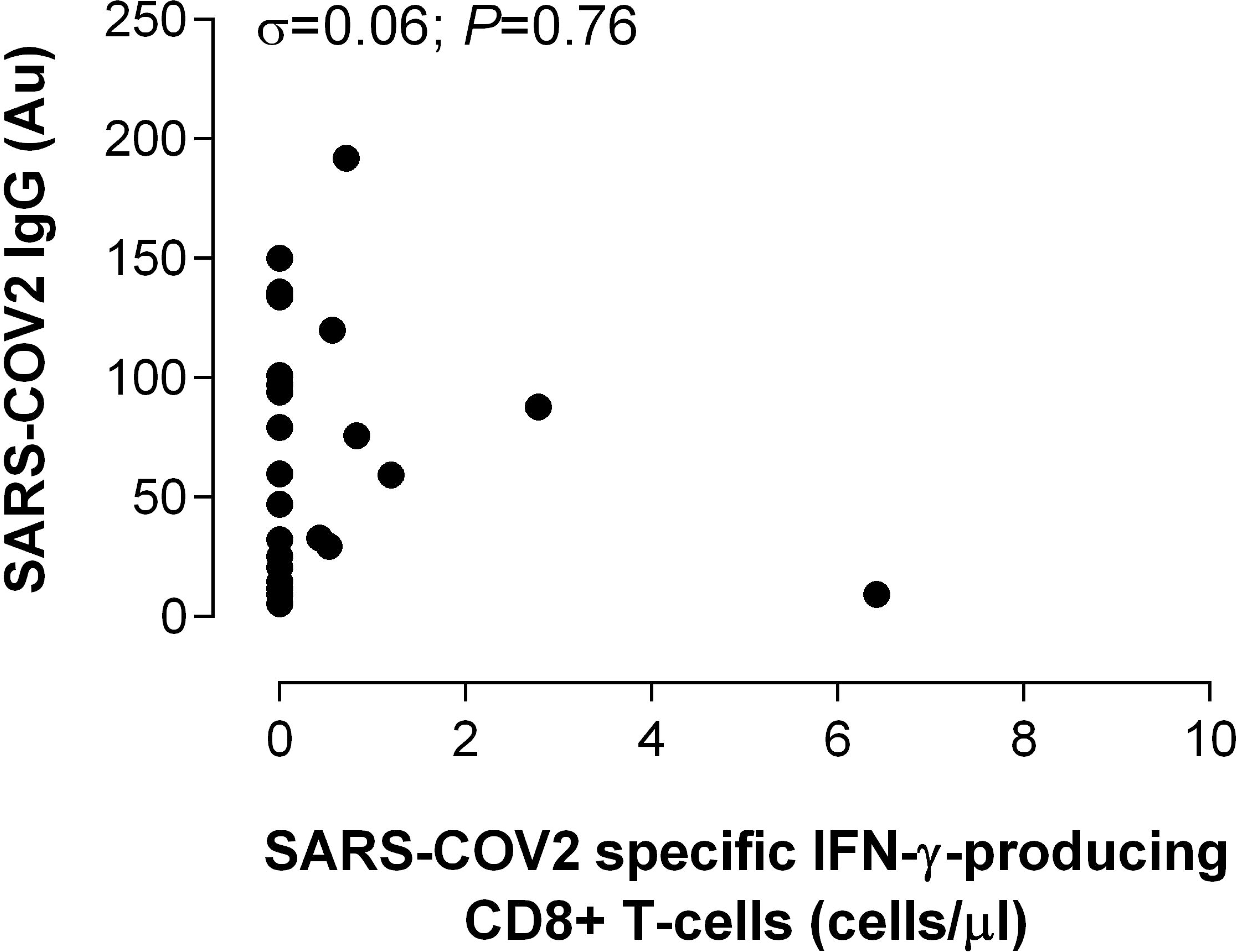
Correlation between SARS-CoV-2-S1/M-reactive CD69^+^-expressing IFN-γ-producing CD8^+^ T cells and serum levels of anti-S-IgG antibodies measured by CLIA. Rho and *P* values are shown.

### Adaptive immune responses to SARS-CoV-2 and laboratory markers of Covid-19 severity

We next investigated whether there was a relationship between SARS-CoV-2-reactive IFN-γ CD8^+^ T-cell counts, S-specific IgG-antibody titers and blood levels of a number of prognostic laboratory parameters of Covid-19 progression, including total leukocyte, lymphocyte and neutrophil counts and markers of inflammation (or coagulation), such as ferritin, CRP and Dimer-D. A pro-inflammatory state was noticed in most patients in this series, while cytopenias were less frequently observed (Table 2). Patients with or without detectable SARS-CoV-2-reactive IFN-γ CD8^+^ T cells were comparable regarding these parameters (Table 2). In addition, no correlation was found between either SARS-CoV-2-reactive IFN-γ CD8^+^ T cells or S-specific IgG-antibody titers and blood cell count or levels of any of these biomarkers (Rho <0.2 and *P*>0.5 in all correlation analyses).

**Table 2.** Laboratory data of patients with and without detectable SARS-CoV-2-reactive IFN-γ-producing CD8^+^T-cells.

| Parameter | Detection of SARS-CoV-2-reactive specific IFN-γ-producing CD8<sup>+</sup> T-cells | | *P* value |
| --- | --- | --- | --- |
|  | Yes | No |  |
| **Days since the onset of symptoms** (median, range) | 32 (11-43) | 26 (2-45) | 0.31 |
| **Total leukocyte count (x10<sup>9</sup>/L)**, (median, range) | 7.41 (5.63-13.58) | 8.76 (4.93-19.80) | 0.53 |
| **Total lymphocyte count (x10<sup>6</sup>/L)**, (median, range) | 1,515 (590-2,910) | 1,360 (200-2,950) | 0.65 |
| **Total neutrophil count (x10<sup>9</sup>/L)**, (median, range) | 5.12 (2.88-10.66) | 5.90 (2.33-19.12) | 0.41 |
| **Ferritin in ng/ml** (median, range) | 428 (61-1079) | 409 (83-1362) | 0.89 |
| **Dimer-D in ng/ml**, (median, range) | 533 (91-1613) | 634 (85-4339) | 0.83 |
| **C-reactive protein in mg/L**, (median, range) | 11 (0.2-124) | 15 (0.7-144) | 0.80 |
| Normal values: 12-300 ng/ml for ferritin, <100 ng/ml for Dimer-D, and <10 mg/L for C-reactive protein. |  |  |  |

## DISCUSSION

There is a knowledge gap regarding the immune mechanisms that confer protection against SARS-CoV-2. On the basis of experimental evidence gathered on SARS-CoV, and MERS-CoV infections,^17,18^ it is assumed that both CD4^+^ and CD8^+^ T cells play a major role in virus clearance and long-term protection against SARS-CoV-2 infection. Although plausible, data supporting this assumption is lacking. In this context, to the best of our knowledge, only two studies have assessed the frequency of SARS-CoV-2-reactive CD4^+^ T (by flow cytometry) or T-cells (by IFNγ-ELISPOT) in peripheral blood from patients with Covid-19 or convalescent individuals, respectively.^13,14^

Here, we optimized a flow cytometry ICS method for quantitation of SARS-CoV-2-reactive-activated (CD69^+^ expressing)-CD8^+^ T cells producing IFN-γ upon antigenic stimulation. This assay uses whole blood as a matrix, thus circumventing the need of PBMC separation, and a combination of two peptide mixes composed of overlapping peptides spanning the S1 region of the S glycoprotein and the entire amino acid sequence of the M protein. We chose SARS-CoV-2 S1 and M proteins as antigenic stimuli because they are expected to contain highly immunogenic CTL epitopes restricted by HLA-class I specificities commonly found in caucasian individuals (all patients in this series), based on sequence alignment with SARS-CoV homologous proteins.^19,20^ Moreover, amino acid sequences of these CTL epitopes are reasonably distinct from aligned sequences present in seasonal coronaviruses, thereby predictably minimizing the likelihood of detecting cross-reactive CD8^+^ T cells.^11^ In turn, by combining S1 and M peptide libraries, we aimed at maximizing our chances of capturing as many SARS-CoV-2 TCR specificities as possible, thus potentially increasing the sensitivity of the assay.

Several findings arose from our study. First, SARS-CoV-2 IFN-γ CD8^+^ T cells targeting S1 and M proteins were detected in 40% of patients at a relatively late time after onset of symptoms (median, 27 days). It is likely that these T-cell subsets could have been circulating long before; our study, however, was not designed to caracterize their kinetics. It is relevant to note that the median age of our patients was high (62 years); although speculative, it can be argued that the rate of detection could have been higher in younger people (no immuosenescence).

Braun and colleagues detected S1 and S2-reactive CD4^+^ T cells expressing cell-surface activation markers in 12 and 15 out of 18 patients (median age, 52 years old), respectively, presenting with mild to severe forms of Covid-19.^13^ The potential antiviral functionality of these T-cell subsets was not explored, though. Ni et al. reported the presence of SARS-CoV-2-specific IFNγ T cells (measured by ELISPOT) targeting the nucleocapsid protein, the M protein or the S1 receptor binding domain (RBD) in up to 7 out of 14 individuals who had recovered from Covid-19.^14^

Second, the frequency of SARS-CoV-2-reactive IFN-γ CD8^+^ T cells varied widely across patients in this series, and was unexpectedly high in some of them (e.g. patient 7 in Table 1:2.75%).

Third, SARS-CoV-2-reactive IFN-γ-CD8^+^ T cells appeared to develop at comparable rates and frequencies irrespective of Covid-19 severity. However, the two patients deceased in this series had no detectable responses. This latter observation is reminiscent of findings reported by Braun et al. who showed that patients with critical disease states lacked S1-reactive CD4^+^ T cells.^13^ In line with this assumption, data published in the SARS-CoV infection setting pointed to an impaired development of adaptive immune responses in patients with severe forms of the disease.^21^

Fourth, interestingly enough, SARS-CoV-2-reactive IFN-γ CD8^+^ T cells were detected in one patient with high clinical suspicion of Covid-19 without microbiological documentation of SARS-CoV-2 infection and testing negative for respiratory viruses and bacteria commonly causing community-acquired respiratory tract infections, thus suggesting that T-cell immunity assays may be an ancillary tool for the diagnosis of Covid-19 in patients with repeat RT-PCR negative testing and delayed antibody conversion.

Fifth, we found no correlation between SARS-CoV-2-reactive IFN-γ CD8^+^ T cells and S-specific antibody levels, which in turn appear to strongly correlate with SARS-CoV-2 neutralizing activity of sera^22^, suggesting that SARS-CoV-2 targeted B and T-cell responses may follow divergent dynamics, as noticed in SARS CoV infection.^19^ Our data is nevertheless in contrast with that reported by Ni et al.^14^ In that study, a strong correlation was found between SARS-CoV-reactive IFNγ T cells and S1-RBD-specific neutralizing antibodies in convalescent individuals (in our current study patients with active disease were included).

Sixth, lung inflammation is the main cause of life-threatening respiratory disorders at the severe stage of Covid-19. It is plausible that SARS-CoV-2-reactive T cells and certain specificities of SARS CoV-2-specific antibodies (i.e. those mediating antibody-dependent enhancement-ADE-) may be mechanistically involved in promoting such a pro-inflammatory state.^23,24^ In this context, we found no correlation between SARS-CoV-2-reactive IFN-γ CD8^+^ T cells, antibodies targeting the S protein (which contain both neutralizing and ADE epitopes) and serum levels of ferritin, CRP and Dimer-D; these findings should not be overinterpreted and by no means rule out the involvement of virus-driven immunopathogenetic mechanisms in progression to acute respiratory distress syndrome. Further prospective studies assessing how dynamics of these parameters relate in sequential specimens are needed to shed some light on this issue.

In addition to the scarce number of patients in our series, the current study has a major limitation: the specificity of SARS-CoV-2-reactive IFNγ-CD8^+^ T cells was not proven. Based on sequence analyses, SARS-CoV-2 and seasonal alpha and beta coronaviruses may share HLA-class I-restricted immunogenic epitopes mapping within S1 and M potentially eliciting cross-reactive T cells.^19^ In support of this assumption, S-reactive CD4^+^ T cells could be detected in 34% of healthy control individuals who had seemingly not been infected by SARS-CoV-2, albeit at lower frequencies compared to patients with Covid-19, and displaying a differential pattern of cell-surface activation markers. We also tested 4 healthy asymptomatic individuals with no evidence of active or past Covid-19 and found one of them to be reactive, albeit displaying a low frequency of SARS-CoV-2 reactive IFNγ-CD8^+^ T cells (0.12%) (data not shown). In an epidemiological framework of heavy SARS CoV-2 community transmission, such as the one being currently faced in Spain, recruiting asymptomatic individuals, even if testing negative by RT-PCR or having no evidence of seroconversion, as negative controls, may not be entirely appropriate, as some of these could have been exposed and developed measurable T-cell responses. Unfortunately, we had not cryopreserved blood specimens from healthy individuals with or without documented infection caused by seasonal coronaviruses collected prior to the penetration of SARS-CoV-2 into our health department. Studies aimed at assessing the specificity of IFNγ CD8^+^ T cells detected by our assay have been designed and planned to be initiated as soon as control of our local epidemic outbreak is achieved.

In summary, in this descriptive, preliminary study we showed that SARS-CoV-2-reactive IFN-γ CD8^+^ T cells can be detected in a non-negligible percentage of patients with moderate to severe forms of Covid-19. As suggested by Braun and colleagues,^13^ either cross-reactive or not, quantitation of these T-cell subsets may provide prognostic information on the clinical course of Covid-19. Studies designed to address this issue are currently underway.

## Data Availability

The data that support the findings of this study are available from the corresponding author,(DN) upon reasonable request.

## ACKNOWLEDGMENTS

No public or private funds were used for the current study. We are grateful to all personnel who work at Clinic University Hospital, in particular to those at Microbiology laboratory for their commitment in the fight against Covid-19.

## AUTHOR CONTRIBUTIONS

EG, EA, IT, MJR and CS, performed T cell and antibody assays, and collected the data. MJG, MLB, MJF, JR and JS-C atended the patients. DN and EG analyzed and interpreted the data and wrote the manuscript.

